# Unique inflammatory profile is associated with higher SARS-CoV-2 acute respiratory distress syndrome (ARDS) mortality

**DOI:** 10.1101/2020.05.21.20051300

**Authors:** Joseph Balnis, Alejandro P. Adam, Amit Chopra, Hau C. Chieng, Lisa A. Drake, Nina Martino, Ramon B. Ramos, Paul J. Feustel, Katherine A. Overmyer, Evgenia Shishkova, Joshua J. Coon, Harold A. Singer, Marc A. Judson, Ariel Jaitovich

## Abstract

The COVID19 pandemic is likely to cause more than a million of deaths worldwide, primarily due to complications from COVID19-associated acute respiratory distress syndrome (ARDS). Controversy surrounds the circulating cytokine/chemokine profile of COVID19-associated ARDS, with some groups suggesting that it is similar to non-COVID19 ARDS patients and others observing substantial differences. Moreover, while a hyperinflammatory phenotype associates with higher mortality in non-COVID19 ARDS, there is little information on the inflammatory landscape’s association with mortality in COVID19 ARDS patients. Even though the circulating leukocytes’ transcriptomic signature has been associated with distinct phenotypes and outcomes in critical illness including ARDS, it is unclear whether the mortality-associated inflammatory mediators from COVID19 patients are transcriptionally regulated in the leukocyte compartment. Here, we conducted a prospective cohort study of 41 mechanically ventilated patients with COVID19 infection using highly calibrated methods to define the levels of plasma cytokines/chemokines and their gene expressions in circulating leukocytes. Plasma IL1RA and IL8 were found positively associated with mortality while RANTES and EGF negatively associated with that outcome. However, the leukocyte gene expression of these proteins had no statistically significant correlation with mortality. These data suggest a unique inflammatory signature associated with severe COVID19.

## Introduction

The COVID19 pandemic has so far caused over seven hundred thousand deaths worldwide, primarily due to complications from acute respiratory distress syndrome (ARDS)^1^. Controversy surrounds the circulating cytokine/chemokine profile of COVID19-associated ARDS, with some groups suggesting that it is similar to non-COVID19 ARDS patients^2^ and others observing substantial differences^3^. Non-COVID ARDS patients who develop a hyperinflammatory signature have a higher mortality than those demonstrating a hypo-inflammatory phenotype; furthermore, these subgroups have different responses to specific interventions such as positive end expiratory pressure (PEEP), administration of simvastatin, and fluid management^4-8^. However, the clinical effects of the inflammatory profile on severe COVID19 infection remains controversial^9^. Moreover, descriptions of the inflammatory signatures in COVID19 patients have been reported using standard clinical laboratory tests^10-16^ or multiplex systems^3,16,17^ that lack adequate methodological calibration of individual cytokines^9,18^. A more precise characterization of circulating cytokine/chemokine expression profile in COVID19-associated ARDS could lead to the development of personalized therapies directed against specific inflammatory signatures^19,20^. Also, because distinct transcriptional signatures in circulating leukocytes have been associated with mortality in critical illness including ARDS^21,22^, it is rational to postulate that COVID19 mortality-associated cytokines are transcriptionally regulated in leukocytes or other sources. Thus, cytokine gene expression levels in circulating leukocytes could provide important insights into the organ-specific regulation of the inflammatory response in COVID19 patients.

In this study, we prospectively interrogated the plasma cytokine/chemokine levels and their leukocyte gene expression in patients with severe COVID19 infection using validated methods. We hypothesized that specific signatures would be associated with mortality in this setting. Portions of this study have been recently reported in a preprint form^23^.

## Methods

We conducted a prospective cohort study involving 41 patients with diagnosis of COVID19 who were admitted to the Albany Medical Center Intensive Care Unit (ICU) and required mechanical ventilation due to ARDS. These patients were enrolled between March 20 and April 17, 2020. Ethical approval was obtained from Albany Medical College Committee on Research Involving Human Subjects (IRB# 5670-20). Patients were considered for enrollment if they were older than 18 years and were admitted to the ICU for invasive mechanical ventilation due to COVID19-associated respiratory failure. Exclusion criteria included likely imminent death or inability to provide consent. Consent was obtained from the patient or a legally authorized representative. Acute illness severity was assessed using the Sequential Organ Failure Assessment (SOFA) score, Simplified Acute Physiology Score (SAPS II) and the Acute Physiology and Chronic Health Evaluation (APACHE II) score^24^. Chronic disease burden information was aggregated with the Charlson Comorbidity index^25^. COVID19 diagnosis was confirmed with regular polymerase chain reaction (PCR) method. Ventilatory system static compliance was obtained at the time of enrollment with patients supported with volume-control ventilatory mode and under full sedation.

### Cytokines measurements

At the time of enrollment, blood samples were obtained and immediately centrifuged for plasma fractionation. Samples were frozen for later cytokine/chemokine determinations using a Human XL Cytokine Magnetic Luminex-Discovery Fixed Panel (R&D Systems #LKTM014) performed simultaneously in duplicates within a week of enrollment completion. Only one freeze-thaw cycle occurred. Cytokines that were found associated with mortality with the multiplex system were later individually measured with enzyme-linked immunosorbent assay (ELISA) using a plate reader (Cytation 5 Imager, BioTek, Winooski, VT), see assays catalogue numbers in **supplemental Table S1**.

### Leukocyte messenger RNA (mRNA) expression determination

In addition to plasma samples, whole blood samples were simultaneously collected from all participants, and leukocytes were isolated using LeukoLOCK^®^ filtering system (catalog no. AM1923; Thermo Fisher). RNA was then extracted from peripheral leukocytes with TRIzol reagent (catalog no. 15596018; Thermo Fisher), and total RNA was isolated following manufacturer’s instructions. Four hundred nanograms of total RNA were used to prepare cDNA using Primescript RT Master Mix at 42°C (catalog no. RR036A; Takara) following manufacturer’s instructions. The cDNA was diluted 10-fold in nuclease-free water, and then, 2μL of cDNA were used per PCR reaction. qPCR was performed in a StepOnePlus (Applied Biosystems) instrument using SYBR green- based iTaq supermix (catalog no. 1725125; Bio-Rad) and 2 pmol primers (Eurofins or Thermo Fisher). Fold induction was calculated using the ΔΔCt method using GAPDH as the reference gene. Each sample was assayed in duplicate and a negative control was included in each experiment. Primer sequences are listed in **supplemental Table S2**.

### Statistical analysis

Due to the long hospital course of severe COVID19 infection^26^, especially in patients receiving mechanical ventilation, the analyzed outcome variable was mortality at day 45 post enrollment. Median (interquartile ranges [IQRs]) and frequency (%) were used to report ICU patient baseline characteristics for continuous and categorical variables, respectively. Associations between mortality and cytokine/chemokine levels obtained with the multiplex Luminex system were analyzed with ANOVA, with *p* value <0.05 considered significant. As multiplex systems require a second inferential step to adjust for false discovery rate (FDR), and because that step can lead to false negative and positive results^18^, any cytokine/chemokine that were found statistically significantly associated with mortality before adjustment for FDR were individually measured with ELISA. For that test, comparison between cytokines/chemokines soluble levels in surviving versus non-surviving patients was accomplished with Student t-test, and a *p* value of <0.05 was considered significant. Association of leukocyte gene products magnitudes in leukocytes with patients’ mortality was done using with Student t-test, and a *p* value <0.05 was considered significant.

## Results

A total of 41 patient were enrolled. The average age of the cohort was 60 years old and there was a significant male sex predominance (68%). 27% were Caucasians, 12% African Americans, 31% Hispanics and 30% other ethnicities. Consistent with previous reports, we found that African Americans and Hispanics had significantly higher mortality than Caucasians^27^. Average APACHE II, SAPS II and SOFA scores were 24, 60 and 9.5 respectively. Other important baseline characteristics are presented in **Table 1**.

**Table 1:**
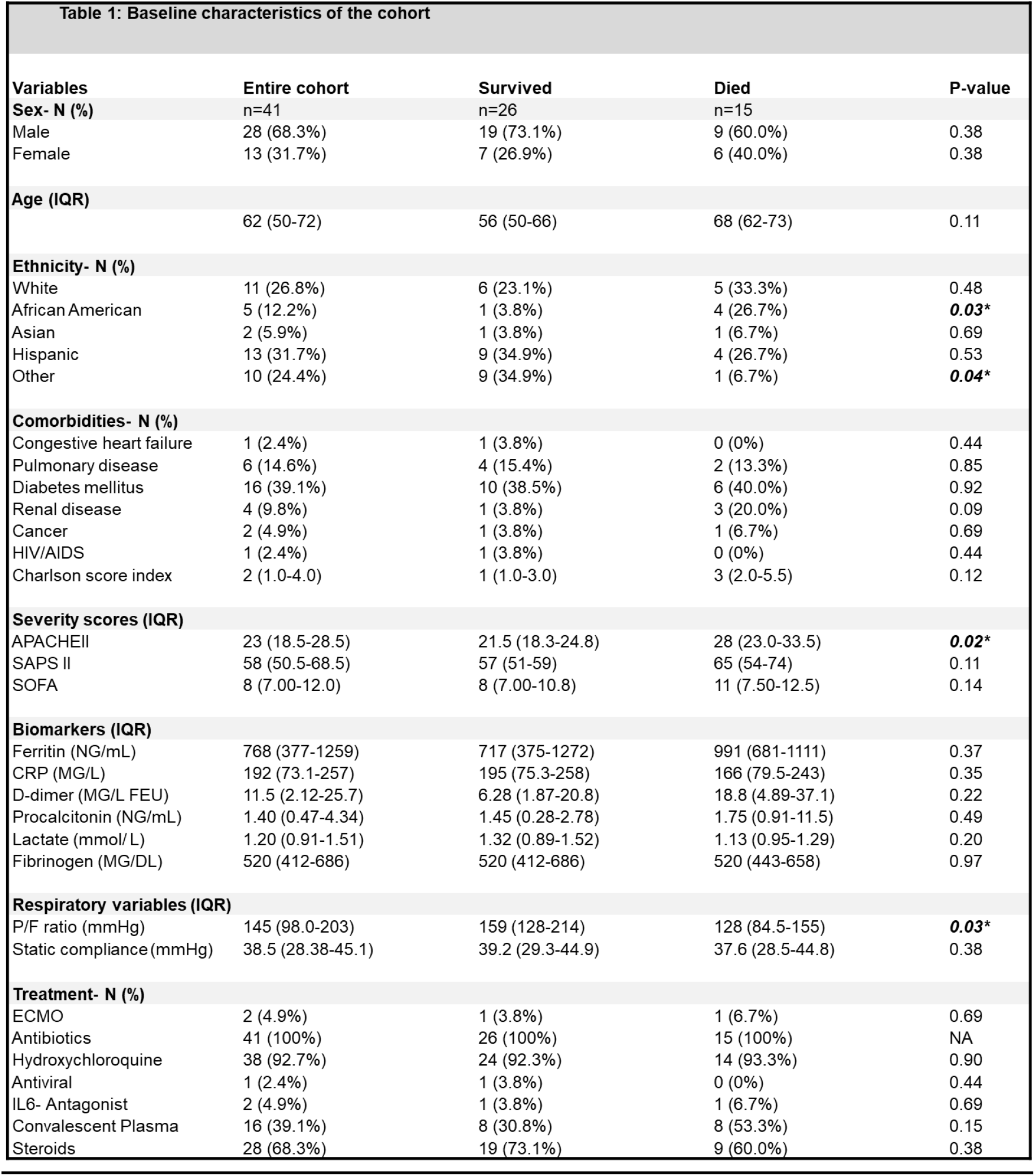
Patients’ baseline characteristics in the whole cohort. Comparison of clinical and laboratory variables between surviving versus non surviving patients. Data are presented as median with interquartile range (IQR); unless otherwise indicated. APACHE II: Acute Physiology and Chronic Health Evaluation II; SAPS II: Simplified Acute Physiology Score; SOFA: Sequential Organ Failure Assessment. ECMO: Extracorporeal membrane oxygenation; CRP: C-reactive protein. Significant *p* values are indicated in bold and italic font. P values for categorical variables are from chi-square tests of association or Fisher’s exact tests when the expected value in any cell is five or less. P values for continuous variables are from Student’s t-test or for non-normal distributed variables by Mann-Whitney rank test.

### Association of cytokine/chemokine circulating levels with mortality

A total of 45 cytokines/chemokines were measured, and 7 were found associated with mortality with the multiplex system, with RANTES, EGF, IL1a and IL5 associated with lower mortality, and IL1RA, IL8 and CCL19 associated with greater mortality (Figure 1-A). The complete dataset including all the cytokines/chemokines, covariables, and outcomes can be found in the https://figshare.com/articles/dataset/Supplemental_file/12808316. IL6, which is associated with worse outcomes in non-COVID19 ARDS, was found unassociated with mortality in this cohort. Out of the 7 cytokines/chemokines that were associated with mortality, individual ELISA determinations confirmed the positive associations of IL1RA and IL8 and negative associations of RANTES and EGF with mortality (**Figure 2**). IL5 and CCL19 levels were unassociated with mortality and IL1a was not detected by the assay (see **supplemental Figure S1**).

**Figure 1:**
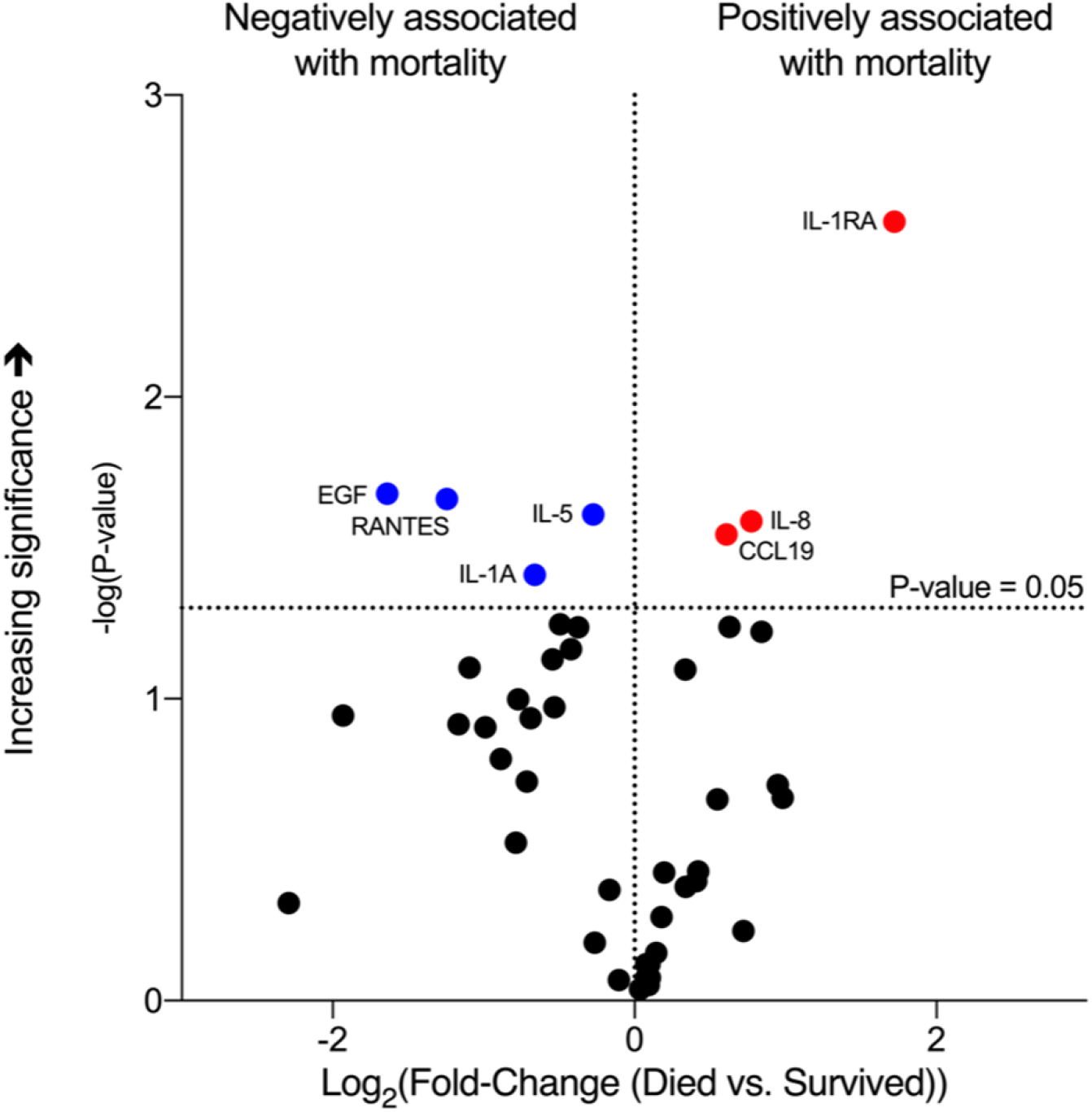
Cytokine determinations at the time of enrollment and association with mortality based on multiplex system determination. Volcano plot showing cytokines/chemokines significantly associated with mortality: blue dots, negatively associated; red dots, positively associated. Black dots identify entities not statistically significantly associated with mortality. Threshold of significance was established at a *p* value of 0.05 before adjustment for false discovery rate.

**Figure 2:**
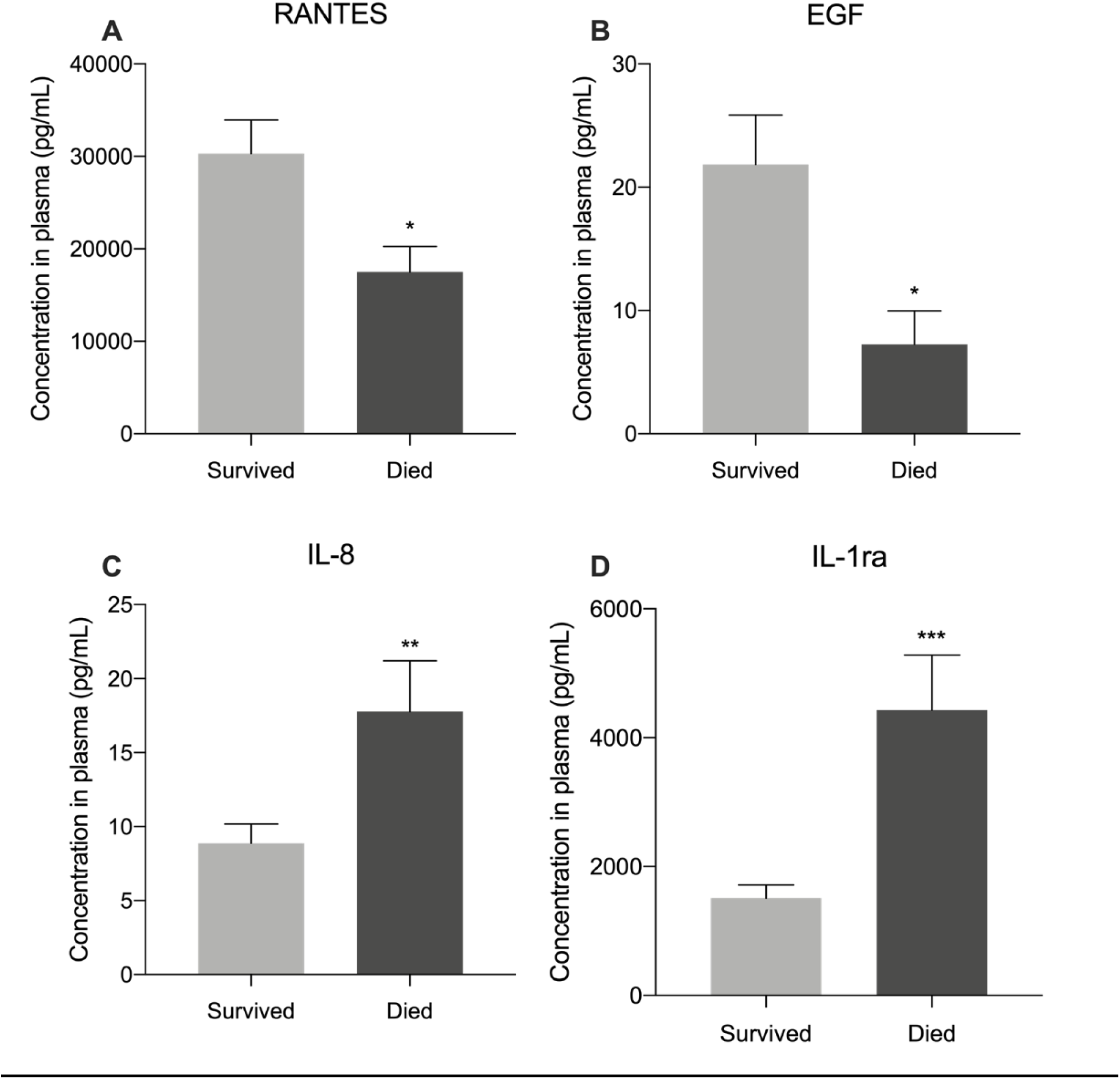
Cytokines associated with mortality based on ELISA tests performed individually. Bar graphs showing the cytokines/chemokines that were found associated with mortality by ELISA testing. \**p*<0.05; \*\**p*<0.01; \*\*\**p*<0.001.

### Association of cytokine/chemokine gene expression levels in circulating leukocytes with their respective protein product and with mortality

Cytokine/chemokine gene expression levels in circulating leukocytes were not significantly associated with mortality. *EGF* and *RANTES* gene expression were modestly but significantly correlated with their respective protein products’ circulating levels, whereas mRNA expression and circulating levels of the other factors were not (**Table 2**).

**Table 2:** Correlation between cytokine/chemokine gene expression in circulating leukocytes with soluble protein and with mortality. Significant *p* values are indicated in bold and italic font.

| Table 2: Correlations with leukocyte mRNA |  |  |  |
| --- | --- | --- | --- |
|  | with soluble protein |  | with mortality |
| Cytokine | R-squared | P-value | P-value |
| RANTES | 0.2318 | **<0.01\*\*** | 0.28 |
| EGF | 0.3277 | **<0.001\*\*\*** | 0.25 |
| IL-1ra | 0.0729 | 0.10 | 0.86 |
| IL-8 | 0.0007 | 0.87 | 0.54 |

## Discussion

We describe a cytokine/chemokine landscape associated with COVID19 mortality, which is different from the one observed in non-COVID19-associated ARDS: Higher IL1RA and IL8, and reduced levels of RANTES and EGF were associated with mortality at 45 days.

The soluble cytokines and chemokines profile associated with COVID19 ARDS and its relationship to mortality has been recently debated and remains incompletely understood^9^. Multiple large ARDS trials have found robust elevations of interleukin (IL)-6^5-7^, and both IL6 and IL8 associate with worse outcomes in that setting^28^. We identified IL8, but not IL6, to be associated with higher mortality in our cohort, suggesting an overlapping but distinct inflammatory signature of COVID19-driven respiratory failure. Consistent with a very recent report^17^, we found IL1 receptor antagonist (IL1RA) to be associated with higher mortality in COVID19. Significantly, while we found that IL1RA levels positively associate with mortality, multiple previous reports indicate that it confers protection against ARDS in non-COVID19 patients. IL1RA is a natural inhibitor of the pro-inflammatory IL1β, which is a highly active cytokine found in the lungs of patients with non-COVID19 ARDS^28-30^. IL1β has been found to increase alveolar permeability both in animals and in humans^31,32^, and experimental models of IL1β-induced lung injury are mitigated by IL1RA administration^33^. Reduced levels of IL1RA in bronchioalveolar lavage samples are associated with higher mortality of ARDS^34^ and IL1RA administration has been proposed as a measure to attenuate lung injury^35,36^. Moreover, a single nucleotide polymorphism (SNP) leading to higher plasma IL1RA levels is associated with reduced ARDS susceptibility^37^. These data suggest that IL1RA elevation represents a feature of COVID19-induced respiratory failure that differs from non- COVID19 ARDS, and the potential pharmacologic modulation of IL1ra has been recently suggested for COVID19 patients^38,39^.

Our finding that RANTES (CCL5) is negatively associated with mortality in our cohort is consistent with a recent report indicating that it may protect COVID19 patients from developing severe disease^3^. RANTES is produced by CD8+ T cells after antigen stimulation and competes with the HIV virus for the CCR5 receptor binding^40^; RANTES could operate in a similar fashion in COVID19 infections. Indeed, a recent study demonstrates that a clonal expansion of CD8+ T cells in broncho-alveolar fluid is associated with milder forms of COVID-19 infection^41^.

There is a growing interest in the value of leukocyte gene expression profiles to define ARDS and sepsis phenotypes and in establishing their association with mortality^22,42^. Indeed, recent data indicates that corticosteroids could be beneficial in certain patient subgroups demonstrating specific leukocyte transcriptomic signatures^19,43^. To gain insight into the regulation of mortality-associated inflammatory mediators in COVID19, we determined the expression levels of the different mortality-associated cytokine and chemokine genes in circulating leukocytes, and found that while some of the interrogated gene expression levels were modestly correlated with the magnitude of circulating soluble protein, none of them were associated with mortality. While future research could elucidate the source of the mortality-associated inflammatory mediators in COVID19, we speculate that the cytokine profile in these patients could be contributed by cells other than circulating leukocytes such as the bronchus-associated lymphoid tissue (BALT)^44^ or by subpopulations of circulating immune cells such as NK cells and others^45^, whose gene expression levels may not be captured with the global leukocyte mRNA analysis. It is also possible that the turnover of these proteins is regulated at the post transcriptional level and thus their expression may not be reflected by the magnitude of mRNA expression.

A main strength of this work is that we describe a prospective cohort of COVID19 ARDS patients and analyzed the association between circulating cytokines and chemokines with mortality. In addition, we determined plasma cytokine levels with ELISA, as opposed to relying on less calibrated clinical laboratory or multiplex testing systems. Indeed, the lack of individual ELISA assays in the current COVID19 literature is considered an important factor that complicates comparing these studies with the non-COVID-associated ARDS trials^9^. Our study has some limitations. First, this is a single-center study that may not be universally reflective of COVID19 ARDS patients. Second, we did not have access to an external validation cohort. However, the associations between IL1RA and worse outcomes and RANTES with better outcomes in COVID-ARDS have been replicated by two recently published studies^3,17^. Third, while we enrolled the patients at the time of hospital admission, we had no control over the period elapsed between the disease development and the blood sampling, which may have affected the inflammatory landscape. Fourth, we had no control of different drugs administered to the patients which could have impacted their inflammatory profile.

## Conclusion

Higher IL1RA and IL8 levels in plasma are positively associated with mortality in a cohort of critically ill COVID19 patients. Elevated RANTES and EGF protein are negatively associated with mortality in these patients. We found no association between gene expression of these cytokines in circulating leukocytes with mortality. While our data indicate that a unique inflammatory signature is associated with severe COVID19 mortality, future research could determine the value of these mediators’ measurements as mortality-predictive biomarkers; and define if their pharmacologic modulation could improve severe COVID19 prognosis.

## Disclosures

None

## Research Support

Part of the results reported herein have been funded by NHLBI of the National Institutes of Health under the award number K01-HL130704 (AJ), and by the Collins Family Foundation Endowment (AJ); NIH/NHLBI 5R01HL049426 (HAS); NIH/National Institute of General Medical Sciences Grant 1R01GM124133 (APA); P41 GM108538 (KAO, ES, JJC).

## Authors Contributions

JB, APA, KAO, ES, JJC, PJF, HAS, MAJ and AJ: Conception and research design, data interpretation and manuscript editing. AC, AJ and HCC: Patients’ enrollments and clinical data collection. APA and NM: multiplex cytokines measurements. JB and LAD: ELISA determinations. RBR: RT-qPCR experiments.

## Supplementary Tables

**Table S1:** ELISA kits

| Target protein | Manufacturer | Product number |
| --- | --- | --- |
| RANTES | Abcam | ab174446 |
| EGF | Abcam | ab217772 |
| CCL19 | R&D Systems | DY361 |
| IL-1ra | Abcam | ab211650 |
| IL-8 | R&D Systems | D8000C |
| IL-1a | Abcam | ab178008 |
| IL-5 | Abcam | ab215536 |

**Table S2:**
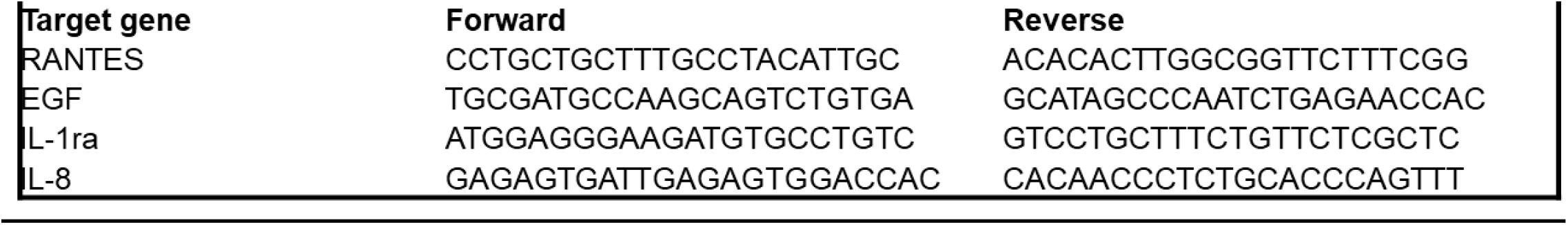
qPCR primers

## Supplementary Figure

**Figure S1.**
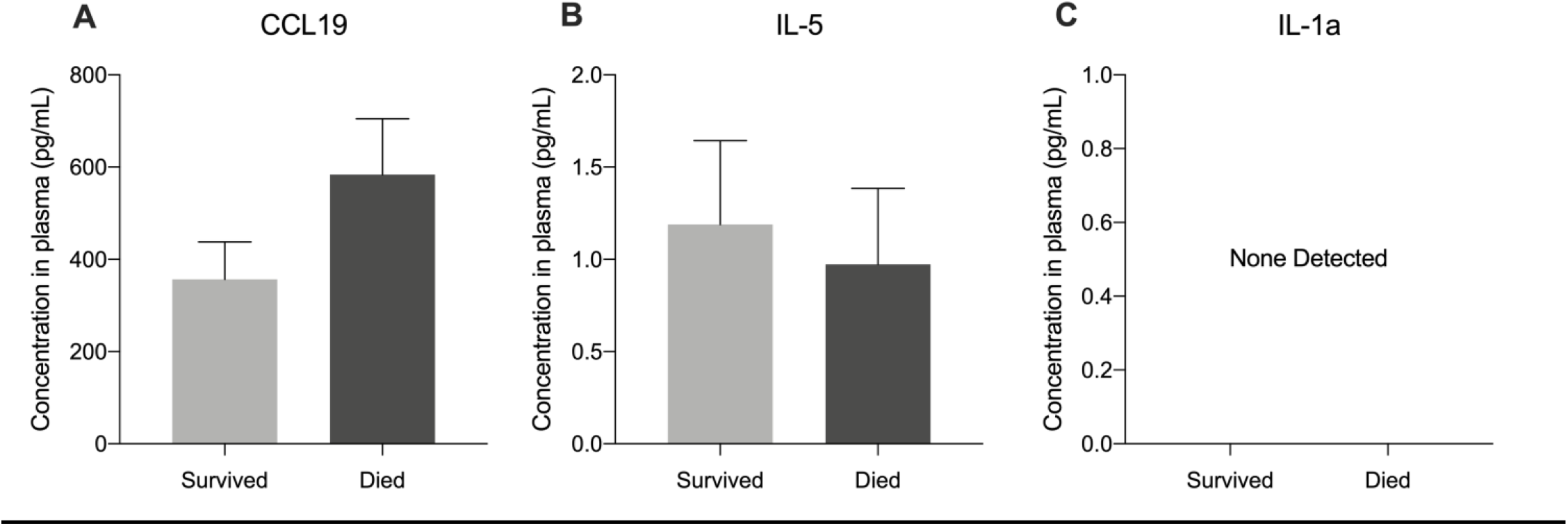

## Data Availability

All the data, including covariables and cytokines are available to the readers

https://figshare.com/articles/dataset/Supplemental_file/12808316

## Notes

### Competing Interest Statement

The authors have declared no competing interest.

### Summary of Updates

Paper is updated based on new data generated with ELISA. The outcome measures was changed to mortality which is more meaningful than liberation from mechanical ventilation, and circulating leukocytes gene transcription was added

